# Barriers and facilitators of implementing a latent tuberculosis infection (LTBI) case management program in a primary care setting

**DOI:** 10.64898/2026.10.23.26366327

**Authors:** Mehabuba Rahman, Rebecca Calderara, Souksavanh Chansombath, John Ho, Masahiro Narita, Yoseph Sorri, Tuberculosis Epidemiologic Studies Consortium

**Affiliations:** Division of Tuberculosis Elimination, Centers for Disease Control and Prevention, Atlanta, Georgia; International Community Health Services, Seattle, Washington; Public Health Seattle and King County, Seattle, Washington

**Keywords:** latent tuberculosis infection, case management, primary care, implementation, qualitative research, barriers, facilitators

## Abstract

**Context:** Approximately 80% of tuberculosis (TB) disease in the United States arises from untreated latent TB infection (LTBI). Primary care clinics are well positioned to address LTBI in populations disproportionately affected by TB, yet implementation barriers remain poorly characterized.

**Objective:** Identify barriers and facilitators encountered during early implementation of a multidisciplinary LTBI case management intervention (CMI) in a federally qualified health center (FQHC) serving predominantly non-U.S.–born patients.

**Design:** Qualitative study using structured key informant interviews, analyzed with iterative thematic analysis. Staff and patients were conveniently sampled; patients were limited to those who completed treatment.

**Setting and Participants:** Three clinics within a FQHC in Seattle, Washington. Thirty-seven interviews were conducted (32 staff, 5 patients) from February 2023-May 2024.

**Main Outcome Measures:** Staff- and patient-reported barriers and facilitators to CMI implementation.

**Results:** Staff identified three barriers: 1) adapting to a new workflow, including technical challenges and role ambiguity; 2) clinic capacity and resource constraints, including staffing and limited language support; and 3) gaps in LTBI care, particularly chest radiograph access and patient questions after a positive test. Staff reported three facilitators: 1) division of labor through role-shifting, enabling detection of side effects and dosing errors; 2) standardized communication tools and educational materials improving consistency and staff confidence; and 3) collaboration via electronic messaging and structured meetings supporting iterative improvement. Patients mostly reported facilitators only, citing the proactive follow-up as central to treatment completion. Both groups cited the drug shortage as a barrier to treatment completion.

**Conclusions:** Multidisciplinary case management revealed facilitators and barriers in delivering coordinated care. Staff identified division of labor, standardized tools, and structured communication key to care coordination while patients emphasized staff encouragement and pro-active follow-up as central to treatment completion. Workflow integration, resource capacity, and post-diagnosis patient support should be considered when implementing or scaling similar models.

## Introduction

Approximately 80% of tuberculosis (TB) disease in the United States arises from untreated latent TB infection (LTBI), a critical and addressable gap in TB prevention.^1^ The United States Preventive Services Task Force recommends LTBI testing and treatment in primary care to reduce TB incidence and improve outcomes across the LTBI care cascade.^2–4^ Non-U.S.–born individuals have an LTBI prevalence 20 times higher than U.S.-born individuals^5^ and face compounding barriers to care, including limited health literacy, financial hardship, and inadequate or absent insurance coverage.^6^ Primary care is a common setting for this population to pursue care, making it an essential touchpoint for TB elimination.^4^

Yet translating screening recommendations into sustained treatment completion remains challenging. Barriers operate at multiple levels: individual factors (limited TB knowledge; side effect concerns); interpersonal factors (patient-provider relationship, cultural concordance of the care team); and health system (short visit times, inadequate interpretation services, insufficient staff training for serving non-U.S. –born patients).^2,7^ Providers report that loss to follow-up and inadequate staffing support, delayed evaluation, and low treatment completion as key challenges.^8,9^ Together, these barriers drive significant patient attrition across the LTBI care cascade.^10^

Multidisciplinary case management may help address these barriers. Case management teams, including nurses, pharmacists, health educators (HEs), and clinicians, can provide patient education in the patient’s preferred language, facilitate follow-up, address side effect concerns, and build the sustained trust needed to support long-term treatment adherence.^7,11–13^ However, evidence on successfully implementing such teams within primary care settings remains limited.

As part of the Tuberculosis Epidemiologic Studies Consortium-III (TBESC-III), International Community Health Services (ICHS) implemented a multidisciplinary LTBI case management intervention (CMI) to support patients across the care cascade, including nurse-led telehealth visits, monthly follow-up calls, care coordination, symptom monitoring, and pharmacy-led medication management. This study explores the barriers and facilitators experienced by staff and patients during early CMI implementation to inform scale-up of similar programs in primary care settings.

## Methods

### Setting and Intervention

ICHS is a federally qualified health center (FQHC) in the Seattle metropolitan area operating 11 clinic sites and serving approximately 36 000 patients annually, the majority of whom are non-U.S.–born. Three ICHS clinic sites participated in the TBESC-III cohort. ICHS serves a patient population that is 76% Asian or Pacific Islander. In 2024, 22% of patients were aged 65 or older, 58% required interpretation services, and 45% lived below 100% of the Federal Poverty Level; 41% were covered by Medicaid, 15% had dual Medicare-Medicaid coverage and 37% were covered by private insurance.^14,15^

The CMI launched in February 2023. The intervention shifted LTBI-related tasks (see Figure 1) across a multidisciplinary team, including an LTBI champion, LTBI registered nurse (RN), HEs (formerly Patient Access Liaisons), and pharmacy staff, to improve LTBI evaluation and treatment completion for non-U.S.–born patients who had not previously completed LTBI or TB treatment. The LTBI champion (R.C.), a provider selected for their LTBI expertise, updated intranet clinical guidelines, led staff trainings and weekly LTBI meetings, and served as the point of contact and expert consultant for ICHS staff, liaising with the county health department. HEs improved electronic health record (EHR)-based patient tracking via standardized templates for follow-up and pharmacy consultations, updated TB/LTBI educational resources, and coordinated ongoing treatment care (e.g., identifying barriers to care, screening for side effects, outreach). The LTBI RN educated patients via telehealth, supported care coordination, and pended select LTBI orders (e.g., labs, medications, chest x-ray). Providers and nurses could quickly order LTBI tests using standardized text shortcuts in the EHR known as “Smart Phrases”. Non-U.S.–born patients were identified using non-English language preference as a proxy. Children under 18 and incarcerated individuals were excluded. All patients with a positive interferon-gamma release assay (IGRA) qualified for case management services.

**Figure 1.**
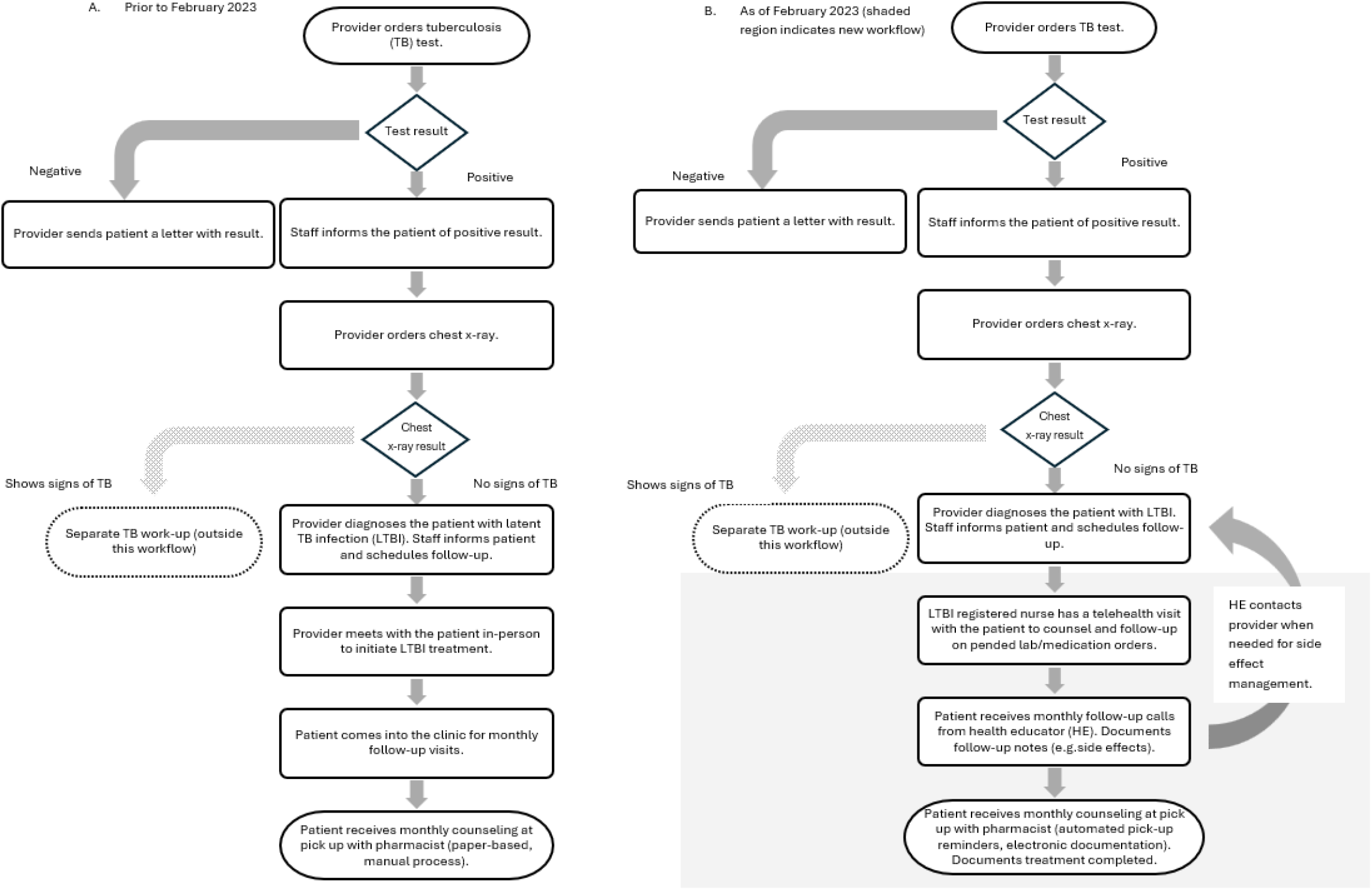
Latent tuberculosis infection clinic workflow before and after case management intervention, 2023.

### Data Collection

Data were collected during the early implementation period when staff were introduced to the intervention and trainings occurred to support acclimation to the CMI. Trainings focused on familiarizing staff with the CMI, including workflow documentation for HEs and LTBI RNs, communication plans, data for the LTBI care cascade, billing, and CMI TBESC-III evaluation metrics. We defined the early implementation period as the time from CMI launch through the point when two consecutive quarters had fewer than two staff trainings, indicating that workflows had stabilized and no further major changes to the intervention were occurring. During this early implementation period, a mean of 10 trainings occurred per quarter.

Structured key informant interviews were conducted quarterly from February 2023 through May 2024. Interviews were led by HEs (S.C.; J.H.) who received training on the interview guides (Appendix A) prior to data collection. HEs had varied educational backgrounds, ranging from associate’s-level training to surgical residency. Interviews were conducted by two female and one male HE. HEs who conducted interviews were also directly involved in delivering the CMI and had an existing care-coordination relationship with many patient and staff participants prior to the interview; participants were aware of the HE’s role in the intervention at the time of interview. Only the HE, interviewee, and interpreter (when needed) were present during interviews; no other individuals attended. Guides were role-specific and received iterative revision. Structured probing questions elicited perceived facilitators and barriers to the early implementation period. Interview duration ranged from 10 to 30 minutes. HEs also maintained memos and debriefing notes throughout data collection, which were reviewed after the final interview. Verbal consent was obtained from all participants prior to interview. Transcripts were not returned to participants for comment or correction.

Staff members were eligible for interviews if they provided direct patient care and interacted with the CMI. Eligible staff were recruited via convenience sampling and invited to participate primarily by email. All invited staff agreed to participate. Four staff members completed two quarterly interviews and one completed four quarterly interviews. Remaining staff completed a single interview. Staff interviews were conducted in English via Google Meet and audio-recorded. Recordings were transcribed using automated Google transcription software, and transcripts were verified for accuracy by HEs. Staff interviews continued until thematic saturation was reached; HEs noted staff theme saturation at 32 interviews. Staff-related findings were reviewed and confirmed by staff involved in CMI implementation.

Patients were eligible for interviews if they completed LTBI treatment while enrolled in the CMI. Eligible patients were recruited by HEs during follow-up calls in the last month of each reporting period (every three months), based on convenience. Unlike staff, no patient was interviewed more than once. Patient interviews were audio-recorded, with telephone interpretation provided in the patient’s preferred language as needed (Mandarin, Cantonese, or Vietnamese). Interviews were transcribed using an encrypted third-party transcription service. HEs concurrently transcribed interpreter-mediated responses within the interview guide. All patient transcripts were verified for accuracy by the interviewing HE. Five (n=5) patients agreed to an interview; no patients declined.

### Analysis

All transcripts were de-identified and analyzed in MAXQDA 24.^16^ A preliminary codebook was developed from 12 de-identified transcripts using a Grounded Theory approach, which emphasized systematic inductive code development, constant comparison, and iterative refinement to ensure analytic rigor.^17,18^ Two HEs (S.C.; J. H.) conducted open coding independently, reconciled discrepancies during coding meetings, and refined codes through successive rounds of 3–4 interviews. A non-ICHS analyst (M.R.) supported the coding process by resolving discrepancies between HE coders and clarifying codebook definitions when needed but did not independently code transcripts. Coders completed the remaining interviews using the finalized codebook and met monthly to discuss discrepancies and code to agreement. Final intercoder percent agreement was 84%. Themes were developed by the non-ICHS analyst and confirmed by staff involved in CMI implementation.

### Human Participant Compliance

This activity was reviewed by the CDC and determined to be not research - program evaluation, conducted consistent with applicable federal law and CDC policy (45 C.F.R. part 46.102[l]). The ICHS research review committee approved the project prior to implementation.

## Results

Twenty-five staff completed 32 interviews. Five patients completed interviews. Among staff interviews, 34% were with providers, 25% HEs, 22% LTBI RNs, and 19% pharmacy staff (Table 1). One-third of interviews (34%; 11/32) involved staff who had worked at the clinic 2 years or less, indicating a substantial proportion of relatively new staff during early CMI implementation.

**Table 1.** Characteristics of staff interviewed about the case management intervention, by role and years at clinic, 2023-24.

|  | By interview n=32 (%)<sup>1</sup> | By unique staff n=25 (%)<sup>1</sup> |
| --- | --- | --- |
| **Role** |  |  |
| Provider | 11 (34) | 11 (44) |
| Health Educator | 8 (25) | 5 (20) |
| LTBI Nurse | 7 (22) | 5 (20) |
| Pharmacy staff | 6 (19) | 4 (16) |
| **Years at clinic<sup>2</sup>** |  |  |
| <1 | 4 (13) | — |
| 1-2 | 7 (22) | — |
| 3-5 | 6 (19) | — |
| 6+ | 2 (6) | — |
| Not reported | 13 (41) | — |

<sup>1</sup> By interview reflects total interviews conducted, including repeats with the same staff member. By unique staff reflects the deduplicated count of individuals.
<sup>2</sup> Years at clinic was not mentioned during the first interviews and an interview question was added in the third quarter to the interview guide. Because years at the clinic was measured at the time of each interview, this value could change for staff who were interviewed more than once during the study period.

Staff identified three primary barriers: (1) adapting to a new workflow, (2) clinic capacity resource constraints, and (3) gaps in care for patients with LTBI (Table 2). Three primary facilitators were identified: (1) division of labor through multidisciplinary role-shifting, (2) standardized communication tools and educational materials, and (3) communication and collaboration. Patients reported mostly only facilitators, describing encouragement and pro-active follow-up as central to their treatment completion. Patients and staff also reported drug shortages as a barrier that temporarily affected treatment completion. Illustrative quotes and themes are seen in Table 2.

**Table 2.** Illustrative quotes from key informant interviews.

| **Barriers** |  |  |
| --- | --- | --- |
| **Theme** | **Subthemes** | **Illustrative quote** |
| Adapting to a new workflow | Increased pharmacy time on treatment documentation | *"Pharmacists are multitasking therefore to pause and do this detailed counseling, is very challenging sometimes. This detailed consultation is not happening in outside pharmacies." – Pharmacy* |
|  | Technical learning curve (Epic-related transition, typing speed, computer skills) | *"There was a learning curve for documenting in Epic for our newer staff and those unfamiliar with Epic and those who have been using Epic but have not checked in patients." – Pharmacy* |
|  | Coordination within clinic | *"It just took a little bit of time to remember what team to task...you're like was it a nurse, was... it care management?" – Provider* |
|  | Intranet as a "culture shift" for accessing new clinical resources | *"Pointing people towards a more aggregated source of information like our clinical intranet, they'd probably use it more, but perhaps it would take a bit of a culture shift as well." - Provider* |
| Clinic capacity and resource constraints | Lack of guidance on non-LTBI resources (e.g. interpretation, CXR access) | *"I've come across more hurdles that I have not had to come across before, so a little bit more direction in where we can go to for resources outside of LTBI." – Health educator (HE)* |
|  | No in-house language support | *"Language barrier has been a very big barrier for me. There are cases, which I just don't know how to help them...if someone who's doing the interpretation for me knows more about, is someone from the organization, who knows the resources better. That could be more helpful." – HE* |
|  | External funding | *"If we could figure out the reimbursement standpoint...it is a time consuming [process]...one of the challenges we had in the past was financial support for programs like this. I am a bit concerned about what happens once this grant is done and where we should get the funding [for] continuing [this work]." – Pharmacist* |
|  | Staffing challenges | *"We do only have two nurses, but we have a lot of patients on our panels. Sometimes they might not get back to a patient." – HE* |
| Gaps in care for patients with LTBI | Cost and insurance coverage | *"I don't know how much exactly a patient is getting the medication, like how much they're paying or how much the copay is." - Provider* |
|  | Transportation to chest x-rays (CXR) | *"I know that we can do free chest x-rays at [County Health Department], but given that a lot of our patients are on the east side, sometimes that is a barrier." – Provider* |
|  | Comprehension of LTBI test results | *"The PCP sent the treatment and by the time they [patient] came to the pharmacy they did not have the registered nurse televisit. So some patients were very confused. Wondering why am I taking this?" - Pharmacy* |
| **Facilitators** |  |  |
| **Theme** | **Subtheme** | **Illustrative quote** |
| Division of labor through multidisciplinary role-shifting | Multiple staff counsel patients and frequent follow-up | *"I think the counseling and the follow up the team provides is incredibly helpful to help getting the patient informed and getting them to make an informed decision to get treatment." - Provider* |
|  | Provider time saved | *"I also really appreciate the support that they provide. It frees up appointment times for providers so that we don't have to do the counseling ourselves." – Provider* |
|  | Providers receive treatment-completion notifications to update records | *"In the past, I wouldn't have known if a patient actually finished treatment, so it's kind of nice that I'm getting that notification and I can actually write it in their chart." – Provider* |
|  | Nurses pend orders (QuantiFERON®-TB (QFT) , medication, CXR), supporting better follow-up | *"I have noticed in the past few weeks that there has been a pended order for a QFT and that has been very helpful. I have really liked that just because it invites a conversation between me and the patient in terms of screening for tuberculosis. All the patients that I talked to so far have been wanting to be tested for TB. The new screening has been doing really well." – Provider* |
|  | Concerns identified: Side effects | *"We were able to catch any gaps, troubleshooting any problem in between for example, if a patient has any side effects, they [HE]* |
|  |  | *usually catch it before the patient actually comes for pick up at the counseling window.” - Pharmacy* |
|  | Concerns identified:<br>Treatment mis-dosing | *“So the patient actually took two pills of the higher dose. At the beginning we did not catch it but [HE] caught it when they asked patient if they were due for next refill and the math was off.” - Pharmacy* |
|  | Concerns identified: Provider prescribing errors | *“There was another case when the patient was prescribed with Rifampin 450 milligram. So that means they take a 150 capsule plus the 300 milligram capsule. There was some confusion with that.” – Pharmacy* |
|  | Concerns identified: Bottled medication dosing errors | *“We’ve identified several patients who despite the counseling were taking the medication incorrectly...even the bottle says something else, translation.” - Pharmacy* |
|  | Concerns identified:<br>Systemic drug shortages | *“So the only issue is I finished the medication on August 20th, but the medication wasn’t available for me to pick up so there was one week that I didn’t have any medication.” - Patient* |
| Standardized communication tools and educational materials | Smart Phrases<sup>2</sup> | *“We have the ‘Smart Phrases’ that we could use as a guide and also save us time when we document, so I think those go pretty well with the intervention.” – LTBI RN* |
|  | Intranet resources | *“I referred to clinical intranet as the next step in this case. There [LTBI Champion] has put in info on TB workup orders.” - Provider* |
|  | Templates | *“I even get educated by the nurse template that they use. I also use that as a reference to remind myself of the side effects as well.” - Provider* |
| Communication and collaboration | Patient outreach | *“In the past three months we’ve been using MyChart<sup>3</sup> which has been helpful and it’s been easier for patients to communicate with us, especially those who might not be available to talk on the phone.” - HE* |
|  | Patient-perceived value of encouragement | *“At the very beginning, I had the nausea along with taking the medication and the nurse calling me saying if you cannot take it, just* |
|  |  | *drop it. But with her encouragement, I am amazed I finished the whole treatment and everything was fine.” - Patient* |
|  | Patient-perceived value of proactive follow-up | *“But when you call and ask me when I have questions I could ask you and that's very reassuring. For example, taking the medicine, two weeks later, my neck has this rash and you helped me schedule an appointment to see the doctor.” - Patient* |
|  | Staff group chat | *“There's also one other thing, the group chat between providers, liaison and the pharmacists. I see a lot of the LTBI liaison using the group chat to ask any questions for drug-drug interaction or medication to hold during the treatment... So it makes a lot of the community communication easy.” - Pharmacy* |
|  | Meetings in person and weekly | *“And also we have the meetings every week, the LTBI meetings weekly to talk about things like, the available workflow or talk about the questions or the different problems we have. And then we try to solve the issues or the problems that we encounter during the LTBI case management. So I think that's great and really helpful to make sure that everything goes smoothly.” – LTBI Nurse* |
|  | Consult resource for complex LTBI cases | *“Love that I can send clinical consults to the group about how to handle unusual cases.” – LTBI RN* |

<sup>1</sup> Order of appearance: Latent tuberculosis (LTBI), Primary care provider (PCP), QuantiFERON® TB-Test (QFT), Chest x-ray (CXR), Tuberculosis (TB), Health Educator (HE), Registered nurse (RN)
<sup>2</sup>Smart Phrases are EPIC templates used to quickly order tests or insert standardized text.
<sup>3</sup>Epic's MyChart is the patient portal used for communication between care teams and patients.

### Barriers

#### Adapting to a New Workflow

Several staff members described the transition to the new CMI workflow as initially disruptive. Pharmacy staff reported that the requirement for detailed written consultations was challenging due to computer malfunctions, the simultaneous transition from NextGen to the Epic EHR system (an event that was not part of the CMI), and the pace of documentation required. This resulted in a steep technical learning curve. One pharmacist noted that patients grew impatient during interactions while documentation was completed. Another acknowledged that while the process was initially “*bumpy*,” it became “*streamlined and smooth*” over time as staff built familiarity and technical skills.

Provider concerns centered on incomplete data transfer between EHR systems and difficulty coordinating (e.g. sending over CXRs) with the county health department to discuss consults, which used a separate EHR. Providers also reported initial ambiguity in coordinating which tasks belonged to the newly onboarded case management team versus the clinical team. Using the intranet as a resource hub represented a “*cultural shift*”, as it had not previously been used for communicating clinical workflows and resources.

#### Clinic Capacity and Resource Constraints

Staff identified systemic resource limitations as a persistent barrier. Staff lacked resources to address non-LTBI barriers that affected treatment adherence such as limited in-house language support. Without language support, patient interactions were lengthy and sometimes ineffective. One HE noted that an in-person interpreter familiar with community resources would have been substantially more helpful than remote interpretation services. Separately, patients and staff also reported a drug shortage, which the team addressed by prioritizing patients at high risk of progression to TB disease and coordinating with pharmacy partners to locate medication.

Even during early implementation, staff raised concerns about long-term funding. A pharmacist noted, *“If we could figure out the reimbursement standpoint…it is a time consuming [process]…one of the challenges we had in the past was financial support for programs like this. I am a bit concerned about what happens once this grant is done and where we should get the funding [for] continuing [this work].*”

Staffing shortages compounded these resource constraints. HE turnover and nursing shortages contributed to staffing gaps. As patient volume grew, staffing remained insufficient to meet increasing case management demands. One HE shared, “*To point out one thing that affects my workflow is the retention of other patient access liaisons. At the moment, I am doing the work alone so I guess that is most important*.”

#### Gaps in care for patients with LTBI

Chest radiograph (CXR) access emerged as a specific and recurring barrier. ICHS clinics did not have on-site CXR services. A CXR result showing no signs of TB is required to rule out TB before starting LTBI treatment. Even when CXR appointments were scheduled, providers and LTBI nurses reported that patients sometimes did not attend due to cost or distance required to reach CXR services. Providers reported that patients who attended the wrong facility received unexpected bills, further undermining trust and follow-up. A provider acknowledged limited visibility into these downstream financial and logistical barriers outside the exam room.

Following a positive TB test, patients often had unresolved questions about next steps and the meaning of the result. This led patients to delay or forgo CXR scheduling. An LTBI RN observed that the opportunity to speak with patients immediately after a positive result, before they were sent for imaging, would help address misunderstandings and improve follow-through.

### Facilitators

#### Division of Labor Through Multidisciplinary Role-Shifting

A central facilitator was the redistribution of LTBI-related tasks across the multidisciplinary team, which reduced time burden on any single staff role and improved continuity of care. LTBI RNs, HEs, and pharmacy staff each provided LTBI counseling, sometimes reinforcing the same education across multiple contacts to improve retention. This allowed providers to focus on higher-complexity tasks such as prescription review, liver function monitoring, and management of concerning side effects. Providers noted that receiving notifications when patients completed treatment, something that did not happen before the CMI, allowed them to update patient records accurately. LTBI RNs took on active roles in following up on pended orders (e.g. QuantiFERON®-TB (QFT), medication, CXR) and explaining normal CXR results in the context of LTBI education.

The multidisciplinary structure also enabled identification of potential problems. During follow-up calls, patients described side effects, such as nausea and discolored urine, to HEs, who provided counseling and connected them with providers when needed. Pharmacy staff identified bottled medications with incorrect dosing milligrams and patient instructions. They launched investigations to correct these errors, which was labor-intensive but critical for accurate treatment completion reporting. Separately, pharmacists observed patients were taking the pills incorrectly and provided re-education. When supply shortages interrupted treatment for one patient, the team identified alternatives through provider coordination and external pharmacy outreach. Previously, these issues often went unnoticed until a patient had already been lost to follow-up.

#### Standardized Communication Tools and Educational Materials

HEs developed documentation templates within Epic for patient follow-up and pharmacy consultations, improving consistency and reducing provider time burden. These templates streamlined discussions about side effects, and one provider noted their usefulness as educational reminders during patient interactions; however, staff suggested the templates could be strengthened by adding more language options for encounters requiring interpretation. “Smart Phrases” in Epic similarly served as a time-saving documentation guide for providers and LTBI RNs. Training materials and intranet resources were described as accessible and confidence-building, enabling clinicians and nurses to offer treatment guidance with greater certainty. The LTBI champion’s intranet summary of guidelines was described by staff as “*quick*,” “*easy to read*,” and “*brief*,” and the intranet also functioned as a centralized hub for other relevant information, such as systemic drug shortages and provider trainings. The value of this training was reflected in one LTBI RN’s account of practice change following a presentation: “*Recommendations from this presentation changed my practice, I recommend screening and treatment to many more patients now than I did in the past*.“

#### Communication and collaboration

Communication adaptations for patient outreach improved patient engagement. Epic’s MyChart messaging and texting for follow-up, adopted after the initial workflow launch, increased responsiveness. According to HEs interviewed, calls placed from a recognizable clinic number seemed to result in more patients picking up.. HEs adapted contact strategies when patients were not reachable by phone. Older patients were especially difficult to contact initially, though HEs reported that establishing a first interaction improved subsequent responsiveness: *“Sometimes I have to call [patients] 5–c times, but once I establish that first interaction… they recognize my number easily and tend to pick up more often.”*

Patients responded positively to the frequency and consistency of check-ins. Patients described staff encouragement and proactive follow-up as instrumental to completing treatment despite early side effects: “*At the very beginning, I had the nausea along with taking the medication and the nurse calling me saying if you cannot take it, just drop it. But with her encouragement, I am amazed I finished the whole treatment and everything was fine*.”

Internal staff communication via MyChart group chat enabled rapid coordination on treatment and follow-up questions. Weekly in-person CMI team meetings provided structured space for problem-solving and allowed staff to voice concerns in a psychologically safe environment: *“No one feels they will be shut down.”* This culture of open communication strengthened team cohesion and enabled iterative improvement to the CMI over time. One provider noted the program was well-regarded enough to prompt discussion of expanding the model to hepatitis B and C management. Providers reported the LTBI Champion was a useful resource for consults, particularly for complex LTBI cases, and was easy to contact given they had EHR-enabled messaging.

## Discussion

This study examined early implementation of a multidisciplinary LTBI CMI within an FQHC serving a predominantly non-U.S. –born, medically underserved population. Findings highlight that successful program implementation requires attention not only to clinical workflows but also to staff technical capacities, organizational communication structures, and health-related social needs, such as CXR imaging access and interpretation, that affect patient engagement.

Several initial barriers, particularly EHR-related documentation challenges, reflected gaps in pre-implementation staff preparation rather than issues in the CMI design. When health systems introduce new programs, conducting a systematic readiness assessment that includes digital literacy and EHR proficiency alongside clinical training may reduce early friction and accelerate workflow stabilization.^19,20^ Similarly, the initial ambiguity about task delegation underscores the value of explicit, documented role definitions at program launch.

Many early barriers were mitigated over time through repetition, structured communication, and organizational flexibility. Notably, communication and documentation initially emerged as barriers but evolved into facilitators as the program matured. Staff described early gaps in role clarity and information-sharing as sources of confusion, while structured communication tools and team meetings became key supports once established. This shift may reflect individual staff adapting over time, differences between staff interviewed earlier versus later, or both. Consistent team meetings with genuine psychological safety for staff feedback were particularly important for iterative program improvement, a finding consistent with implementation science and case management literature emphasizing the role of organizational learning and role redistribution in successful scale-up.^7,11–13,21^ However, this organizational learning depends on staff continuity, and the high turnover and understaffing noted above may limit the extent to which such gains are sustained as the program scales.

Patient-level barriers, including cost, transportation, and test interpretation confusion, following a positive TB test result reflect well-documented challenges in LTBI care cascades^10^ that might be amplified in low-income, non-U.S.–born populations.^7^ The CMI team’s care coordination role helped address some of these health-related social needs, but resource constraints limited the team’s capacity to fully resolve barriers such as insurance coverage gaps and transportation. Staff raised concerns about sustainable reimbursement of externally-funded programs.

Notably, the five patients interviewed among those who completed treatment raised these barriers, which may reflect selection bias, patient self-efficacy, or a supportive case management team. Although the sample was small and limited to those who completed treatment (n=5), this absence may suggest that proactive CMI follow-up and care coordination mitigated common obstacles. Patient accounts among treatment completers consistently emphasized facilitators, aligning with literature on the role of social support in LTBI treatment completion.^22^

### Implications for Policy & Practice

- **Prepare staff before launch.** New programs should assess non-clinical skills (e.g., EHR proficiency, documentation speed) alongside clinical training. Ramp-up time and training can reduce workflow friction and role ambiguity.
- **Invest in standardized communication tools and structures early.** Documentation templates, EHR-embedded quick phrases, and shared intranet resources built before implementation improved consistency and staff confidence.
- **Build deliberate patient and staff communication channels.** Multi-channel outreach (calls, texting, MyChart) and persistent recontact may have likely enhanced patient responsiveness; internal group chats and accessible consult channels may support problem-solving and team cohesion.
- **Prioritize case management team sustainability from the beginning**. Division of labor across LTBI RNs, HEs, and pharmacy staff supported detection of side effects before patients were lost to follow up and dosing errors while reducing burden on any single role. Turnover and reimbursement concerns threatened sustainability.
- **Address structural and linguistic barriers proactively.** Staff identified recurring barriers, including CXR access and limited language support. Programs should consider on-site imaging partnerships and interpretation during intervention development.
- **Close communication gaps at key decision points**. Providers reported that patients often had unresolved questions after a positive TB test, delaying radiograph follow-through. Connecting patients with staff immediately after a positive result may improve completion.

### Limitations

This study has several limitations. HEs delivered the intervention and conducted interviews from a convenience sample, introducing potential social desirability and confirmation bias. Mitigation strategies included the use of two independent coders and a non-ICHS analyst, application of a Grounded Theory approach with its systematic coding procedures, and iterative refinement of the codebook to agreement. Additionally, because some staff were interviewed across multiple quarters, the 32 staff interviews reflect fewer than 32 unique individuals, which may affect the diversity of perspectives captured relative to the total interview count. Patient perspectives were limited by the small sample size (n=5), which may not have been sufficient to reach thematic saturation, and interviews were restricted to patients who completed treatment. Therefore, patient perspectives should be interpreted as illustrative rather than generalizable. Barriers faced by patients who did not complete treatment are unknown and the absence of saturated patient-reported barriers may reflect this selection bias rather than true saturation of such barriers. Quantitative measures of the intervention’s effectiveness should be assessed given the strong, positive qualitative feedback reported in this study. Findings reflect one FQHC in the Pacific Northwest and may not generalize to primary care settings serving different populations or operating under different policy and resource conditions.

## Conclusion

Implementing a multidisciplinary LTBI CMI in a primary care setting is feasible but requires deliberate preparation, standardized communication tools, and ongoing role clarification. Early barriers related to workflow adaptation, resource constraints, and gaps in LTBI-specific follow-up co-occurred with facilitators such as division of labor, standardized documentation and educational materials, and consistent team and patient communication. Patients reported that staff encouragement and proactive follow-up supported their treatment completion, though exploration into LTBI treatment barriers is needed for patients who did not complete treatment. Staff perceived that shifting LTBI treatment initiation visits from providers to specialized LTBI RNs through virtual care models, along with pharmacist-led monitoring for medication side effects and case management services provided by trained HEs, reduced provider time burden and supported more patient-centered care. These findings offer practical lessons for health systems seeking to implement or scale multidisciplinary, telehealth-enabled LTBI case management, including the importance of pre-launch technical readiness, standardized communication tools, and proactive support for patients navigating post-diagnosis care. Future research should evaluate whether implementation processes translate into measurable improvements in treatment completion, clinical outcomes, and cost-effectiveness across diverse primary care settings.

## Data Availability

Deidentified individual participant data that support the findings of this study may be available from the International Community Health Services (Rebecca Calderara) upon reasonable request after approval of a proposal.

## Acknowledgments

We would like to acknowledge the contributions of Zahra Masroori, ICHS staff and patients, and Kathryn Winglee to this manuscript.

## Disclaimers

All parts of this manuscript were refined using Claude Sonnet 5 to revise text to improve flow and grammar and authors take responsibility for the integrity of all content.

The findings and conclusions in this report are those of the authors and do not necessarily represent the views of the Centers for Disease Control and Prevention. References in this manuscript to any specific commercial products, process, service, manufacturer, or company does not constitute its endorsement or recommendation by the U.S. Government or CDC.

## APPENDIX A

### Key Informant Interview Questions

Thank you for agreeing to participate in this interview. I work at International Community Health Services (ICHS). I am conducting this interview as part of the Tuberculosis Epidemiologic Studies Consortium (TBESC) III program. TBESC is a CDC-funded initiative to carry out research on latent (or sleeping) tuberculosis. ICHS has partnered with the CDC to help conduct this research. The interview will be used as part of our evaluation plan to identify what is going well and what needs improvement in our treatment of latent tuberculosis. You are being asked to take part in this study because you are a provider who has had LTBI patients.

You may not benefit directly from the study. However, your responses may help us learn how to improve latent tuberculosis care at ICHS. Participation is completely voluntary, and will only be needed once for this interview. No identifying information will be collected, and any identifying information in your answers will be removed. The information provided will remain strictly confidential. You may choose not to answer any question.

**Do you have any questions before we get started?**

**Do I have your permission to begin the interview?**

**Do I have your permission to record the interview?**

**2 people giving interviews-1 giving questions/1 recording verbatim (or recording s transcribing)**

**Beginning**

- For interviewer

- Who is the interviewer?
- Background of interviewer?
- For interviewee

- How long have you been at ICHS?
- How much interaction do you have with the intervention?

#### Intervention: CASE MANAGEMENT

##### PROVIDER

**What is going well with the case management intervention?**

Probes:

- Have you sent a patient for case management services? If yes, did the patient have any questions?
- Do you think case management is improving LTBI treatment adherence? Why or why not?

**What is not going well with the case management intervention?**

Probes:

- What are challenges or barriers to the program?
- How could the program be improved?

##### NURSE

**What is going well with the case management intervention?**

Probes:

- Do you believe the initial telehealth nurse visits have been helpful for patients? Why or why not?
- Have you been able to provide increased LTBI patient education to patients through the case management program? If yes, what was helpful? If not, what was not helpful?
- Do you think case management is improving LTBI treatment adherence? Why or why not?

**What is not going well with the case management intervention?**

Probes:

- What are challenges or barriers to the program?
- How could the program be improved?

##### PHARMACY

**What is going well with the case management intervention?**

Probes:

- In your role as a pharmacist, have you collaborated with the case management program to support patients with LTBI in medication adherence? How has your experience been?
- Have you been able to provide increased LTBI patient education to patients through the case management program? Any specific experiences/examples you’d like to share?
- Do you think case management is improving LTBI treatment adherence? Why or why not?

**What is not going well with the case management intervention?**

Probes:

- What are challenges or barriers to the program?
- How could the program be improved?

##### Health Educator

**What is going well with the case management intervention?**

Probes:

- Which Patient Access Liaisons methods do you think have been the most beneficial or effective in ensuring treatment adherence among LTBI patients?
- Do you think case management is improving LTBI treatment adherence? Why or why not?

**What is not going well with the case management intervention?**

Probes:

- What are challenges or barriers to the program? For instance has patient care been difficult with the refilling process or any other case management process?
- How could the program be improved?
- From your interactions with LTBI patients, what are the main challenges or barriers that they face in adhering to their treatment plans?

##### PATIENTS

**What is going well with the case management intervention?**

Probes:

- Did you find case management helpful or effective in your LTBI care? If so, which aspect of it? (initial encounter, symptom check-ups, med-refills, etc)
- Do you think case management is improving LTBI treatment adherence? Why or why not?

**What is not going well with the case management intervention?**

Probes:

- What are challenges or barriers to the program?
- How could the program be improved?
- Is there anything you wish could have been done differently in terms of the support or assistance received from case management?

##### END

Did you have any other questions or comments that have not been addressed that you would like to bring up?

